# Prioritizing embryos with lower homozygosity is expected to reduce disease risk in children of related individuals undergoing preimplantation genetic testing

**DOI:** 10.64898/2026.05.30.26354526

**Authors:** Tobias Wolfram, Mohammad Ahangari, Ivan Davidson, Laura Wartschinski, Jeremiah H. Li, Mathew Eyre, David Stern, Justin Schleede, Alireza Haghighi, Shai Carmi, Michael Christensen

**Affiliations:** Herasight Research, USA; Harvard Medical School, Boston, MA, USA; Department of Medicine, Brigham and Women’s Hospital, Boston, MA, USA; The Broad Institute, Cambridge, MA, USA; Braun School of Public Health and Community Medicine, The Hebrew University of Jerusalem, Israel

**Keywords:** autozygosity, runs of homozygosity (ROH), consanguinity, PGT-A, PGT-M, embryo selection, recessive disorders, congenital anomalies, neurodevelopmental disorders

## Abstract

Consanguinity is a reproductive union between individuals who share a recent common ancestor. These unions are common in many regions of the world and increase the burden of rare recessive disorders by elevating autozygosity in offspring. Current reproductive genetic screening focuses on a limited set of known pathogenic variants, leaving most recessive risk unaddressed. Here we argue that embryo-level autozygosity, quantified as the fraction of the genome in long runs of homozygosity (*F*_ROH_), is a potentially actionable genomic biomarker that can be integrated into routine preimplantation genetic testing as a homozygosity-informed embryo-prioritization framework (PGT-H) that can be layered onto existing embryo biopsy workflows when couples are already undergoing IVF with PGT-A or PGT-M. Using forward simulations of first-cousin and double-first-cousin couples, we show that siblings conceived by the same couple span a wide range of *F*_ROH_; selecting the lowest-*F*_ROH_ candidate from a cohort of five embryos reduces *F*_ROH_ by approximately 40% on average. Combining these reductions with empirical effect-size estimates, we estimate that for first-cousin couples this strategy could reduce risk of intellectual disability by roughly 35 – 45% (corresponding to an absolute risk reduction of about 1.8 – 2.2%) and potentially reduce excess recessive disease burden, while also modestly reducing risk of common diseases such as type 2 diabetes. We outline how existing PGT-A and PGT-M workflows could potentially be extended to report embryo-level *F*_ROH_ and discuss ethical and counseling considerations. Autozygosity-based embryo prioritization offers a principled way to address a component of recessive risk that current variant-centric approaches miss.

## Introduction

Consanguinity is a reproductive union between individuals who share a recent common ancestor (Figure 1A). Consanguineous marriage is common in many parts of the world (Figure 1B) and can confer social and economic benefits ^1^. Accordingly, reproductive counseling in this context should remain culturally sensitive and non-directive. However, it is well established that offspring of related parents are at elevated risk for various poor health outcomes ^2–7^. When parents share recent ancestors, they may transmit long genomic segments that are identical by descent. This results in elevated genome-wide autozygosity in their children, that is, genomic regions inherited identically by descent from both parents. Elevated autozygosity increases the chance that rare recessive variants are homozygous in the offspring, thereby conferring elevated disease risk. This mechanism underlies the long-recognized excess of recessive disease in consanguineous families.

**Figure 1:**
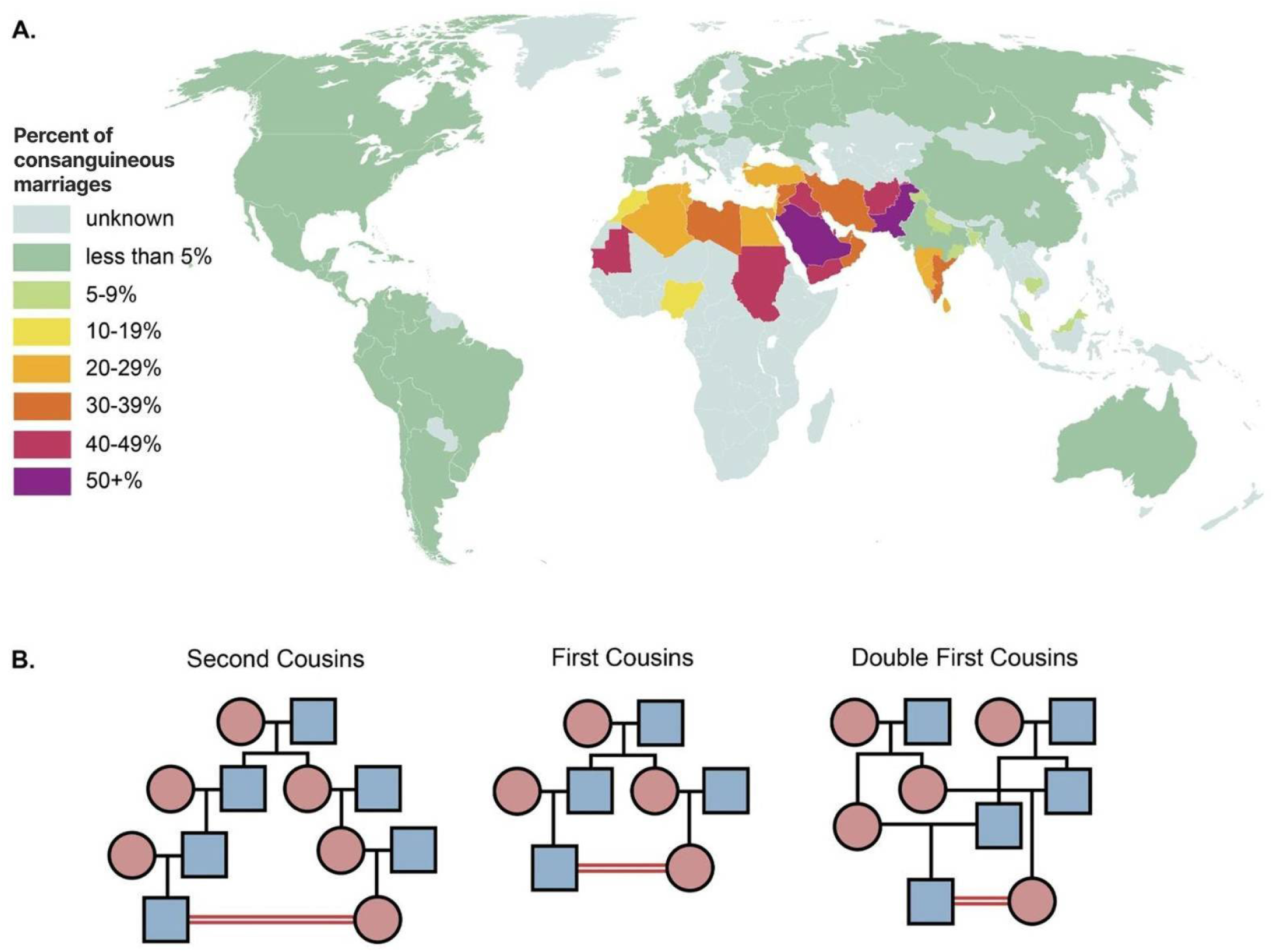
Consanguinity and its global distribution. (A)Estimated proportion of marriages between second cousins or closer by country, adapted from Hamamy et al. ^9^ (B)Pedigrees illustrating second-cousin, first-cousin, and double-first-cousin unions (red double lines indicate the consanguineous partnership). These relationships yield progressively higher expected offspring autozygosity (expected F_ROH_ = 1.5625%, 6.25%, and 12.5%, respectively, in the absence of background homozygosity).

Autozygosity can be measured from runs of homozygosity (ROH), which represent long homozygous genomic segments inherited identically by descent from both parents (Figure 2). In outbred populations, the fraction of the genome in ROH, or *F*_ROH_, has values usually below 1%, whereas offspring of first-cousin unions have an average ROH of 6.25%. Offspring of double first-cousin unions have an average ROH of 12.5%. Individual *F*_ROH_ values, however, can vary substantially even among full siblings due to Mendelian transmission and the stochastic nature of meiotic recombination. This within-family variation creates a clinical opportunity to preferentially select embryos with lower autozygosity even when parents have a fixed degree of relatedness.

**Figure 2:**
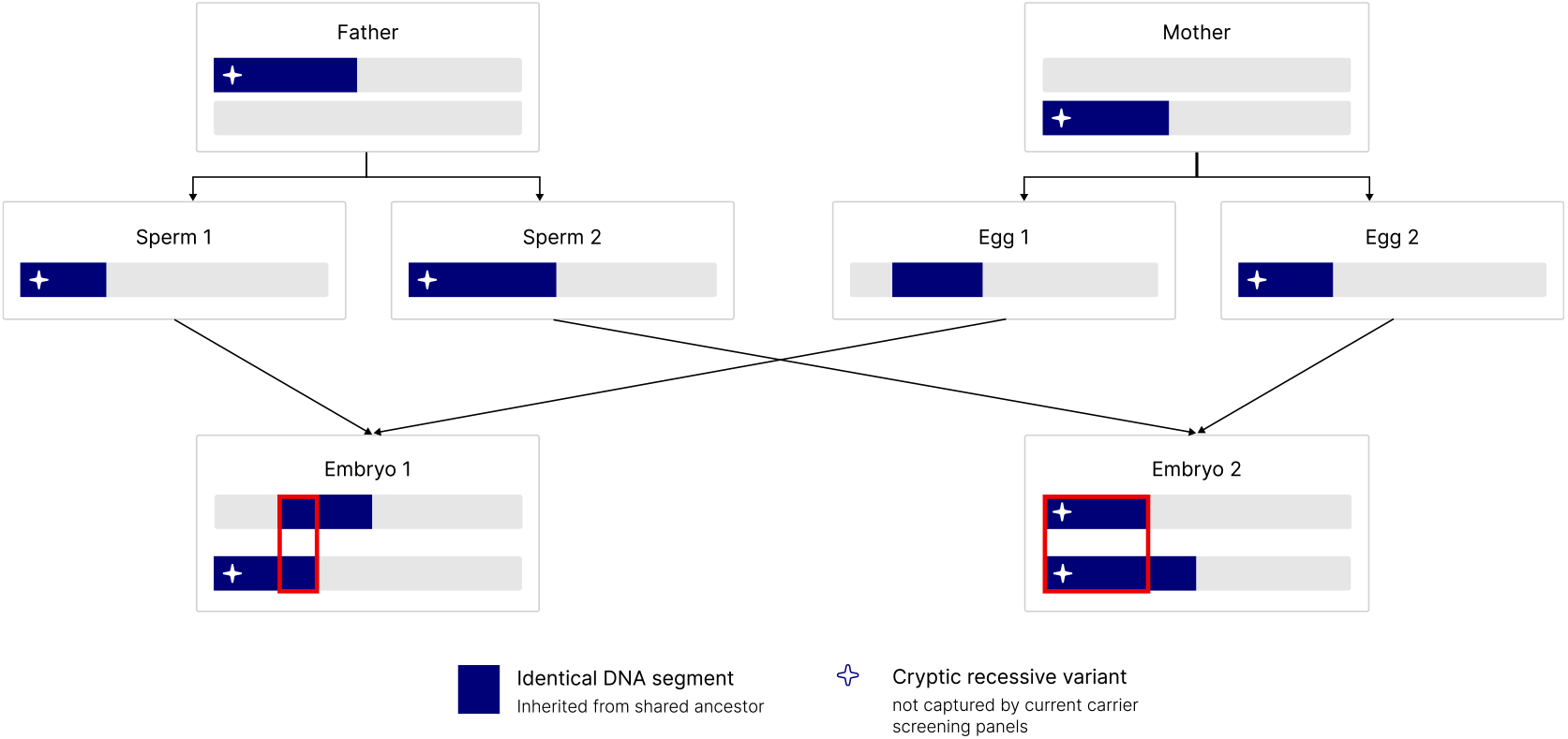
Autozygosity and health outcomes. Schematic showing how having shared recent ancestors generate long identical-by-descent segments (runs of homozygosity, ROH) in offspring; recombination and meiotic division produces embryo-to-embryo variation in the location and extent of ROH and can render recessive variants homozygous (crosses), including previously unidentified or unscreened variants not currently captured by standard carrier panels.

For reproductive medicine clinicians, the practical question is how to reduce the genetic risks (including both Mendelian and complex polygenic architectures) posed by elevated autozygosity in consanguineous couples. Long-standing ethical analyses stress that it is inconsistent with genetic counseling principles to discourage consanguineous marriage ^8,9^; rather, the emphasis should be on communicating evidence-based facts for informed decision-making and providing access to medical resources such as genetic testing, when applicable. In this article, we synthesize clinically relevant evidence, explain why offspring *F*_ROH_ varies widely within families and describe the utility of prioritizing embryos with lower realized autozygosity through preimplantation genetic testing for homozygosity (PGT-H), as well as how this framework can be integrated into standard IVF workflows. We emphasize what is necessary for responsible clinical adoption and realistic counseling about risk reduction rather than risk elimination.

### Elevated autozygosity is associated with increased risk of rare disorders and common disease-related outcomes

The best-established consequence of elevated autozygosity is an increased burden of autosomal recessive disease. This has been demonstrated through population-genetic modeling, autozygome studies, and sequencing-based analyses ^10^, and several large-scale studies in populations with high rates of consanguinity have shown that a meaningful fraction of pediatric severe intellectual disability, multiple congenital anomalies, and other rare disorders can be attributed to autosomal recessive variants that are homozygous in regions of ROH ^11–14^.

Beyond classic Mendelian conditions, genome-wide autozygosity has been associated with a broad range of common traits and diseases. Analyses in diverse cohorts have linked higher *F*_ROH_ to increased risk of cardiometabolic disease, lower height, reduced cognitive performance, adverse reproductive outcomes, higher BMI, increased risk of type 2 diabetes, and reduced fertility or increased risk of childlessness ^2–5^. These associations are typically modest at the individual level but can translate into substantial population-level burdens in areas with high consanguinity.

Epidemiologic studies of consanguineous unions provide convergent evidence. Relative to offspring of unrelated parents, children of first-cousin or more closely related couples have increased risk of stillbirth, infant death, congenital anomalies, and developmental delay ^7,15–17^. In a Norwegian registry study spanning over two decades, consanguinity was associated with elevated rates of stillbirth and infant mortality even after adjusting for maternal education and other covariates ^15^. In the Born in Bradford cohort, children of first-cousin parents had elevated risks of mortality, higher healthcare utilization, and increased rates of speech, language and learning difficulties, with corresponding impacts on educational outcomes ^16^.

Taken together, these findings demonstrate that elevated autozygosity is a biologically interpretable and clinically meaningful risk factor for a mixture of monogenic and polygenic outcomes. Importantly, *F*_ROH_ captures the genome-wide burden of recessive variants, including those at loci not yet known to be associated with disease. This motivates embryo selection strategies that directly target autozygosity rather than only known variant-level risk.

### Large within-family variation in autozygosity among embryos enables substantial disease risk reduction

Although parental relatedness determines the expected level of autozygosity in offspring, the realized *F*_ROH_ for any given child depends on the stochastic details of recombination and Mendelian segregation. Even among full siblings from the same consanguineous couple, *F*_ROH_ can vary widely. Embryo screening can leverage this within-family variation by prioritizing the embryos with lowest *F*_ROH_.

To quantify this variation, we performed forward simulations of consanguineous pedigrees using realistic sex-specific recombination maps ^18^. For each relationship type (first cousins and double first cousins), we simulated 1,000 independent families. For each simulated couple, we generated 1,000 embryos by sampling sex-specific recombination events along the parental haplotypes and tracking the inheritance of founder chromosomes through the pedigree. ROH segments were defined as autosomal tracts where the two homologous chromosomes were inherited from the same founder haplotype, and *F*_ROH_ was computed as total autosomal ROH length divided by total autosomal genome length.

Within each family, we characterized the distribution of embryo *F*_ROH_ by computing the family mean, the standard deviation, and the range across 1,000 embryos. As expected, mean *F*_ROH_ clustered near the theoretical values of 0.0625 for first-cousin unions and 0.125 for double-first-cousin unions, but individual embryos spanned a broad range around these means. Figure 3A illustrates the distribution of mean *F*_ROH_ across simulated first-cousin families, and Figure 3B shows the distribution across embryos for a representative family, highlighting substantial within-family variance.

**Figure 3:**
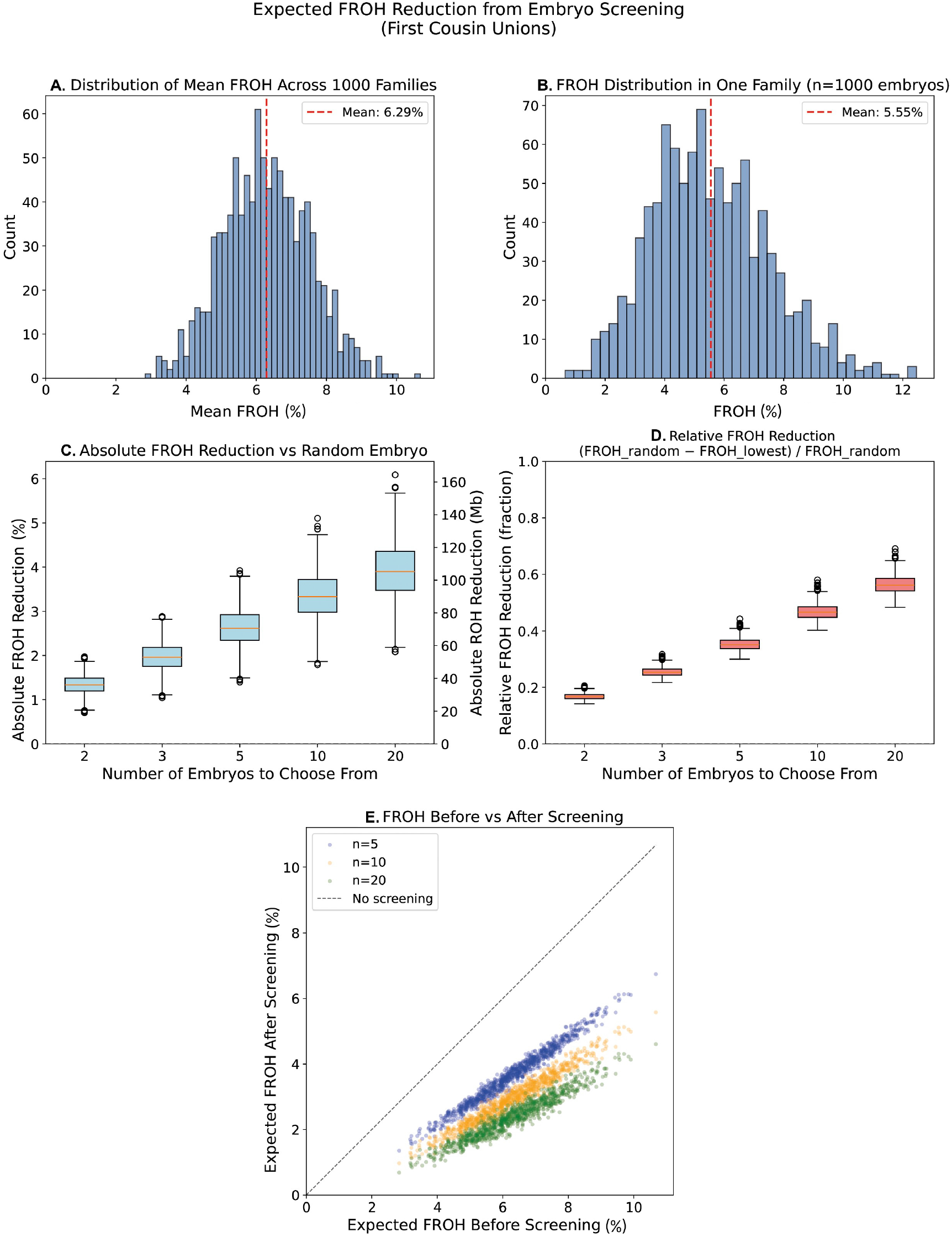
Simulated expected reduction in embryo *F*_ROH_ from screening in first-cousin unions. (A) Distribution of mean F_ROH_ per family across 1,000 simulated first-cousin families. (B) Distribution of F_ROH_ among 1,000 embryos from a representative family. (C) Expected absolute reduction in F_ROH_ comparing random embryo to lowest-F_ROH_ embryo from sets of n = 2, 3, 5, 10, 20 embryos. (D) Same as (C) showing relative risk reduction. (E) Expected embryo F_ROH_ with versus without screening. Each point represents a simulated family. For each family, the expected F_ROH_ is calculated as the mean embryo F_ROH_ across 1,000 simulations when embryos are selected at random versus when embryos are prioritized by F_ROH_ from a cohort of n embryos.

We then asked how much autozygosity reduction could be achieved by selecting the embryo with lowest *F*_ROH_ from clinically realistic cohort sizes. For each family and each embryo count ranging from 2 to 20, we repeatedly sampled a given number of embryos without replacement from the pool of 1,000 embryos and recorded the minimum, mean, and maximum *F*_ROH_ across 10,000 replicates. Importantly, our simulations assumed no background *F*_ROH_ in the population, which may underestimate the absolute reduction achievable, especially in populations with elevated background autozygosity. Because background autozygosity may increase within-family variance in some populations, our simulations may underestimate absolute selection benefit in those settings; however, the magnitude of this effect requires population-specific validation. To corroborate the simulation results, we also developed a mathematical model to approximate the reduction in *F*_ROH_ as a function of the number of embryos (see Supplement).

For first-cousin couples, selecting the lowest-*F*_ROH_ embryo from five embryos reduced realized *F*_ROH_ by an average of roughly 40% relative to a randomly selected embryo, with larger gains as the number of embryos increased (Figure 3C, Figure 3D, Supplementary Figure 1). For double-first-cousin couples, absolute *F*_ROH_ reductions were higher but the proportional reductions from embryo selection were smaller (average reduction of 3.8 *F*_ROH_ % points, meaning a relative reduction of approximately 30%).

Finally, comparing expected *F*_ROH_ with and without screening demonstrated consistent improvement across all families, with larger embryo cohorts yielding lower *F*_ROH_ (Figure 3E).

We emphasize that the above analysis assumes that all considered embryos have birth potential. Thus, the relevant parameter is the number of potential *births* per IVF cycle, rather than the number of embryos ^19^.

### Translating reductions in autozygosity into expected health benefits

Translating reductions in autozygosity into the full extent of health-related benefits is challenging because autozygosity has been associated with dozens of diseases and traits, and the extent to which each of these associations is biologically causal remains unclear in the absence of within-sibling replications. Rather than attempting to model all such outcomes, we focus on a small set of well-established and high-impact outcomes with evidence supporting plausible recessive contributions: infant mortality, intellectual disability, congenital anomalies, type 2 diabetes, and childlessness. The first three are likely driven predominantly by rare, large-effect recessive variants, whereas the latter two likely reflect diffuse recessive contributions from many loci.

For outcomes where the increased risk is likely due to single large-effect recessive variants, we assume that the excess risk above the outbred baseline scales approximately linearly with *F*_ROH_. For an individual with *F*_ROH_ = *F* and a deleterious recessive variant of frequency *p*, the probability of homozygosity is approximately *pF* for small *p*, so doubling *F* roughly doubles the contribution of such variants to disease risk. In a two-step screening framework, PGT-M first removes a fraction of that risk by targeting known pathogenic variants. For common, polygenic outcomes such as type 2 diabetes and childlessness, we instead model the log-odds of disease as proportional to *F*_ROH_, as has been empirically observed in within-family analyses that help minimize environmental confounding ^3–5^. In both cases, the key intuition is that lowering *F*_ROH_ reduces the chance that recessive-acting alleles are homozygous across the genome, so that each percentage-point reduction in autozygous genome content translates into a proportional reduc-tion in excess recessive disease risk.

We combined published estimates of baseline risks in outbred populations with relative risks as a function of parental relatedness (via *F*_ROH_) to estimate absolute risks for first-cousin and double-first-cousin couples with and without embryo screening (see Methods for details). For intellectual disability and congenital anomalies, we further incorporated the expected diagnostic yield of exome-based testing to model the fraction of recessive risk that could be removed by variant-level PGT-M alone, ^12,20^ versus the additional reduction achieved by autozygosity-based embryo selection. We note that this likely provides an overly optimistic estimate for the effectiveness of PGT-M because PGT-M is typically restricted to pathogenic or likely pathogenic variants, whereas clinical diagnoses in affected individuals include variants that would not meet this classification threshold. For intellectual disability and congenital anomalies, we assume the baseline attributable fraction from recessive-acting variants is 4%, since empirical estimates suggest biallelic variation accounts for around 4% of neurodevelopmental disorders in outbred populations ^21^, and assume the same value for congenital conditions since no precise estimates of the attributable fraction are known to our knowledge. Although we also modeled infant death as having a primarily monogenic genetic architecture, we could not identify sufficiently relevant estimates for the expected PGT-M risk reduction. Finally, we attribute the excess risk associated with increased *F*_ROH_ in these conditions entirely to the elevated burden of monogenic-acting homozygous recessive genotypes.

These estimates depend on simplifying assumptions regarding the linearity of recessive burden, diagnostic yield, and the relationship between *F*_ROH_ and disease risk. Table 1 summarizes the resulting expected risk reductions for the two relationship types. For double first-cousin couples, the combination of PGT-M and *F*_ROH_-informed embryo screening removes about half of the excess risk for the primarily Mendelian outcomes we considered. For intellectual disability in double first-cousin couples, the absolute risk decreases from 8% to 5 – 6.2% with PGT-M alone (assuming 30 – 50% diagnostic yield ^13,20,22^) and to 4.1 – 4.9% with the combined strategy.

**Table 1:**
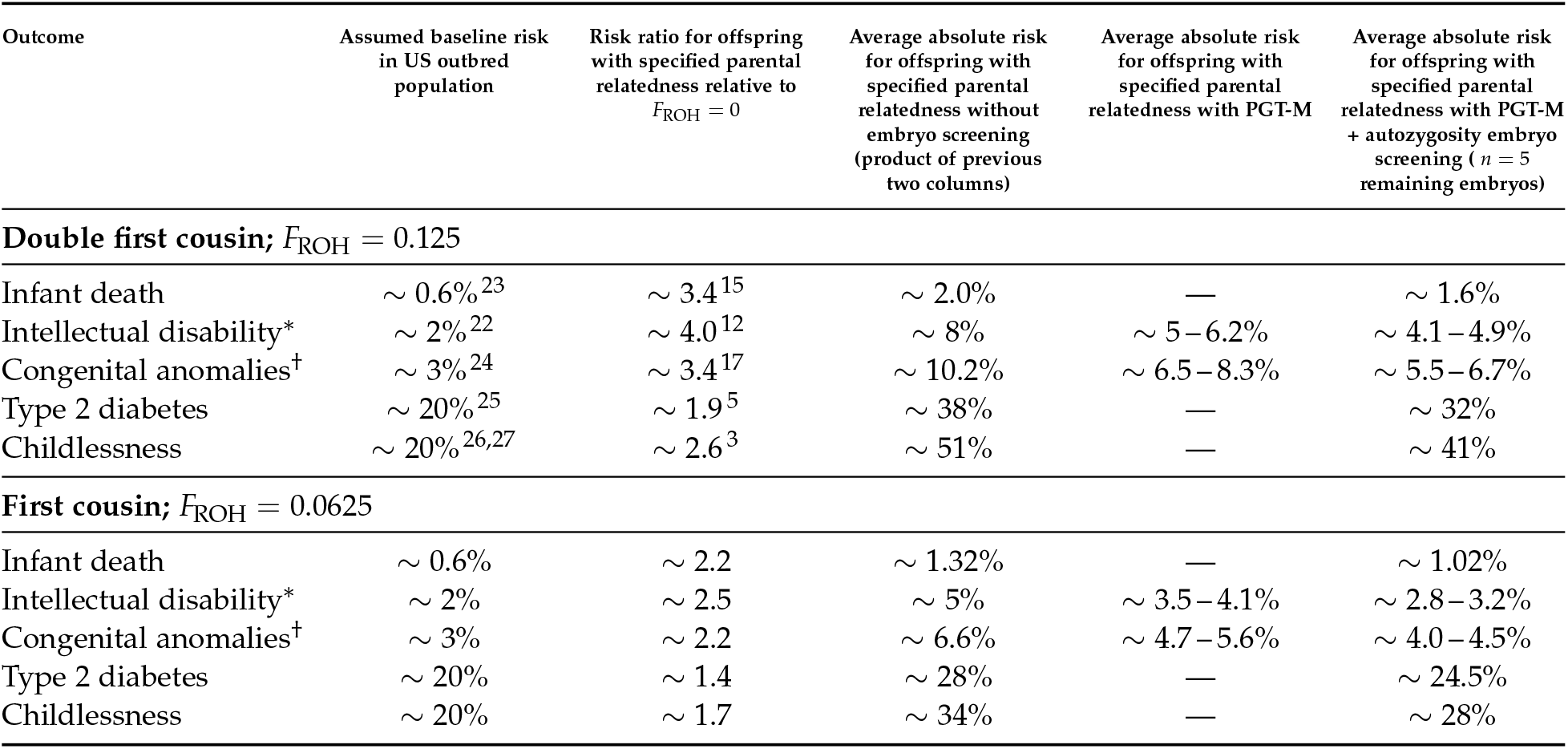
Expected risk reductions after screening for autozygosity. For infant death, congenital anomalies, and intellectual disability, where the increased risk is likely due to large-effect recessive variants, we assume the risk ratio varies linearly with F_ROH_. For type 2 diabetes and childlessness, we assume log-odds vary. linearly with F_ROH_. See Methods for details. ^*^For intellectual disability rows, PGT-M values assume a 30 – 50% diagnostic yield ^13,20,22^. ^†^For congenital anomaly rows, PGT-M values assume a 26 – 50% diagnostic yield ^20,28^

| Outcome | Assumed baseline risk in US outbred population | Risk ratio for offspring with specified parental relatedness relative to $F_{ROH} = 0$ | Average absolute risk for offspring with specified parental relatedness without embryo screening (product of previous two columns) | Average absolute risk for offspring with specified parental relatedness with PGT-M | Average absolute risk for offspring with specified parental relatedness with PGT-M + autozygosity embryo screening ($n = 5$ remaining embryos) |
| --- | --- | --- | --- | --- | --- |
| **Double first cousin; $F_{ROH} = 0.125$** |  |  |  |  |  |
| Infant death | $\sim 0.6\%$<sup>23</sup> | $\sim 3.4$<sup>15</sup> | $\sim 2.0\%$ | — | $\sim 1.6\%$ |
| Intellectual disability* | $\sim 2\%$<sup>22</sup> | $\sim 4.0$<sup>12</sup> | $\sim 8\%$ | $\sim 5-6.2\%$ | $\sim 4.1-4.9\%$ |
| Congenital anomalies<sup>†</sup> | $\sim 3\%$<sup>24</sup> | $\sim 3.4$<sup>17</sup> | $\sim 10.2\%$ | $\sim 6.5-8.3\%$ | $\sim 5.5-6.7\%$ |
| Type 2 diabetes | $\sim 20\%$<sup>25</sup> | $\sim 1.9$<sup>5</sup> | $\sim 38\%$ | — | $\sim 32\%$ |
| Childlessness | $\sim 20\%$<sup>26,27</sup> | $\sim 2.6$<sup>3</sup> | $\sim 51\%$ | — | $\sim 41\%$ |
| **First cousin; $F_{ROH} = 0.0625$** |  |  |  |  |  |
| Infant death | $\sim 0.6\%$ | $\sim 2.2$ | $\sim 1.32\%$ | — | $\sim 1.02\%$ |
| Intellectual disability* | $\sim 2\%$ | $\sim 2.5$ | $\sim 5\%$ | $\sim 3.5-4.1\%$ | $\sim 2.8-3.2\%$ |
| Congenital anomalies<sup>†</sup> | $\sim 3\%$ | $\sim 2.2$ | $\sim 6.6\%$ | $\sim 4.7-5.6\%$ | $\sim 4.0-4.5\%$ |
| Type 2 diabetes | $\sim 20\%$ | $\sim 1.4$ | $\sim 28\%$ | — | $\sim 24.5\%$ |
| Childlessness | $\sim 20\%$ | $\sim 1.7$ | $\sim 34\%$ | — | $\sim 28\%$ |

We estimated intellectual disability risk of about 5% without screening for first-cousin couples, compared with around 2% in the outbred population ^12,13^. Under the same assumptions, PGT-M alone reduced this to roughly 3.5 – 4.1%, and PGT-M plus *F*_ROH_-based embryo screening reduced it further to 2.8 – 3.2% when five embryos remain after PGT-M. Infant death risk is reduced from approximately 1.3% to 1.0% when embryos are prioritized on *F*_ROH_, and type 2 diabetes risk decreases from about 28% to 24.5%. These estimates illustrate that PGT-M and *F*_ROH_-based embryo screening act in a complementary fashion: PGT-M removes risk mediated by known variants, whereas *F*_ROH_-based prioritization reduces the remaining genome-wide recessive burden in proportion to the achievable reduction in embryo-level autozygosity. It should be emphasized that these risk reductions are achieved simultaneously for all of these and other conditions. This differs from polygenic embryo screening approaches, where prioritizing embryos specifically for low risk for one condition may not reduce risk for other genetically distinct conditions.

### Clinical implementation and counseling considerations

Consanguinity arises in diverse cultural, religious and socio-economic contexts, and a consanguineous marriage often provides social and economic benefits ^1,9^. Ethical analyses and professional guidelines emphasize that discouraging consanguineous marriage per se is inconsistent with core principles of genetic counseling, which prioritize autonomy, non-directiveness, and respect for cultural values ^8,29–33^. Instead, the focus should be on communicating evidence-based information about risks and options, and on ensuring equitable access to testing and reproductive technologies. Ethical arguments developed around polygenic embryo screening provide a useful parallel framework for thinking through disclosure, counseling, and permissible uses of embryo autozygosity screening ^34^.

Within this framework, *F*_ROH_-based embryo prioritization must be presented as a risk-reduction tool, rather than a guarantee against disease. Counseling should avoid framing PGT-H as a recommendation against consanguinity and should instead present it as an optional risk-reduction tool within a non-directive reproductive counseling framework. Counseling should clearly distinguish between: (i) risks from known pathogenic variants that can be targeted by PGT-M; the genome-wide, largely uncharacterized recessive risk reflected in *F*_ROH_; and (iii) other genetic and non-genetic contributors to adverse outcomes. Counseling should also emphasize that autozygosity-informed selection typically reduces relative risk by tens of percent, but leaves a substantial residual risk due to remaining ROH, non-recessive etiologies, and unscreened genetic mechanisms. It should also be emphasized that the reduction in ROH can vary substantially across families and embryo cohorts for a given family. Finally, it is important to communicate that risk reductions are likely to be smaller than what may be estimated for the given number of embryos, as not all embryos will result in a live birth, and some embryos may be carriers of monogenic pathogenic variants. More formal counseling aids that account for embryo attrition and cohort size can help translate embryo-level ranking into realistic expectations of benefit at the live-birth level ^35^.

In practice, preconception or early-pregnancy genetic counseling for consanguineous couples should include discussion of the couple’s genealogical relatedness, expected *F*_ROH_ range in off-spring, and how PGT-A, PGT-M, and PGT-H intersect. For couples already undergoing IVF with PGT-M or PGT-A, embryo genotyping or sequencing data generated in standard workflows can support estimation of embryo-level *F*_ROH_, particularly when combined with parental genotype data or haplotype information (Figure 4). Recent work has shown that embryo genomes can be reconstructed from ultra-low-pass PGT-A sequencing using parental genomic data, supporting the feasibility of deriving additional embryo-level genomic measures from existing PGT data ^36^.

**Figure 4:**
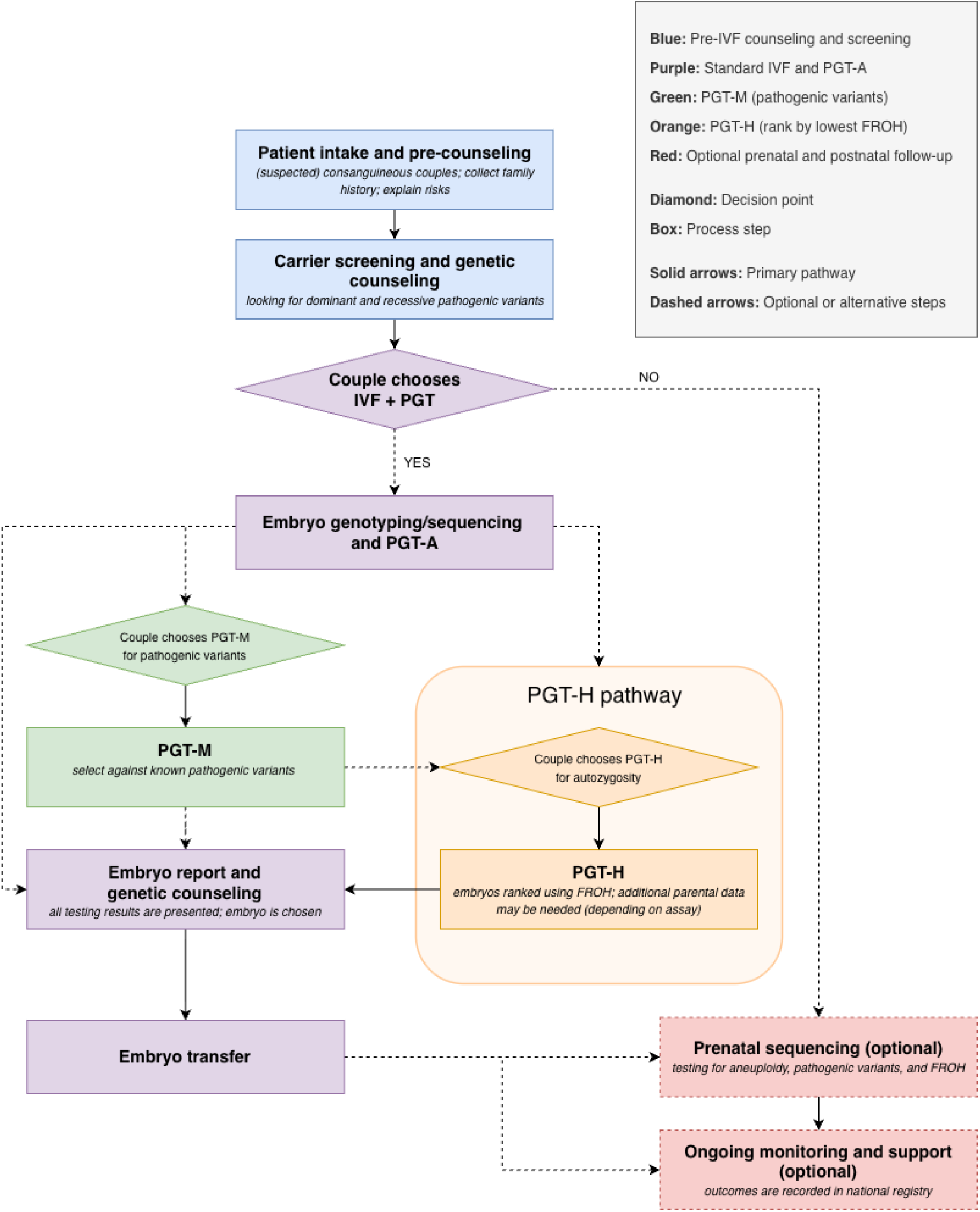
Workflow for integrating *F*_ROH_ into routine PGT-A / PGT-M. Parental sequencing or genotyping enables construction of chromosome-scale haplotypes. Ultra-low-pass sequencing of embryo biopsies is already performed for PGT-A, from which embryo-level F_ROH_ can be computed and reported alongside aneuploidy and monogenic findings. The workflow illustrates how F_ROH_-informed embryo prioritization (PGT-H) can be layered onto existing pre-IVF counseling and screening, carrier screening and genetic counseling, standard IVF with PGT-A and/or PGT-M, embryo transfer, and optional prenatal sequencing and follow-up for couples already undergoing IVF with embryo biopsy.

When parental genotype data are available, as is typical when carrier screening is performed, ROH segments identified in embryos can be interpreted in light of the stated pedigree and parental haplotype structure, helping distinguish expected consanguinity-derived autozygosity from atypical homozygosity patterns.^1^

In cycles yielding multiple euploid embryos, counseling may need to aid couples’ interpretation and decision-making when multiple distinct metrics are presented, such as expected *F*_ROH_ reduction and relative and absolute risk reduction for early and late onset conditions.

Importantly, expanding access to genomic testing and PGT must not exacerbate health inequities. Consanguinity is more prevalent in populations that already face structural barriers in healthcare and research representation ^9,16^. Ensuring fair access to PGT-H will require efforts in insurance coverage, culturally sensitive outreach, and development of infrastructure in regions where consanguinity is common.

## Conclusion

Genome-wide autozygosity is a robust, biologically interpretable risk factor that links the long-known epidemiology of consanguinity with modern genomic data. Our synthesis of the literature supports the conclusion that elevated *F*_ROH_ increases risk for a spectrum of rare and common outcomes, and our simulations show that siblings conceived by the same highly consanguineous couple can differ markedly in *F*_ROH_. Selecting embryos with lower autozygosity therefore offers a principled way to reduce recessive disease burden beyond what is achievable by variant-centric PGT-M alone.

*F*_ROH_-based embryo prioritization can be implemented with data already generated in routine PGT-A and PGT-M workflows. We estimate that for first-cousin and double-first-cousin couples, PGT-H in combination with PGT-M can reduce absolute risk of severe outcomes such as intellectual disability and major congenital anomalies by several percentage points, and reduce excess risks (compared to unrelated couples) by roughly one-half to two-thirds.

These risk reductions are meaningful but they do not prevent disease, which must be communicated as such. PGT-H should be framed as one tool among many for consanguineous families, complementing carrier screening, variant-level PGT, and prenatal diagnosis. Several limitations should be considered. Our modeling assumptions push our estimates in a conservative direction: Background autozygosity of 1-3%, common in populations with historically elevated consanguinity, ^3^ would widen within-family variance and increase the absolute benefit of selection. Diagnostic yields in affected individuals may likely overstate what PGT-M can prevent in practice, since variants identified as diagnostic in affected probands would not necessarily meet clinical thresholds for PGT-M. We anticipate that autozygosity-based embryo selection may eventually become integrated into reproductive genomics in settings where consanguinity is common.

## Methods

### Simulation of embryo-level autozygosity in consanguineous families

We simulated consanguineous pedigrees to model the distribution of embryo-level *F*_ROH_ for first cousin and double-first-cousin couples. Founder haplotypes were generated on an idealized autosomal genome. Crossovers were modeled as a Poisson process along each chromosome, without interference, and recombination rates were determined by the sex-specific map for each parent ^18^.

For each relationship type, we simulated 1,000 families. In each family, we drew founder haplotypes, propagated them forward through the consanguineous pedigree by sampling crossovers and segregating chromosomes at each meiosis, and produced 1,000 embryos for the final couple. Rather than simulating full genotypes, we traced the boundaries of founder haplotypes as they were transmitted across generations. For every embryo we recorded, for each autosomal chromosome, the piecewise-constant haplotype identity inherited on each chromosome. We then constructed the union of all breakpoints from the two homologous chromosomes of each embryo, partitioning each chromosome into minimal contiguous intervals over which haplotype identity was constant. An interval was classified as ROH (homozygous-by-descent) if the two haplotype IDs were identical, indicating that both copies traced back to the same founder haplotype. The physical lengths of all autosomal ROH segments were summed to obtain total ROH length per embryo, and *F*_ROH_, which is expressed as a percentage, was computed as total autosomal ROH length divided by total autosomal genome length.

Within each family, we used the 1,000 simulated embryos to characterize the underlying *F*_ROH_ distribution. We estimated the family-specific mean *F*_ROH_, which corresponds to random embryo choice, and the variance and range across embryos. To model embryo selection, we considered embryo cohort sizes *n* ∈{2, 3, 5, 10, 20}. For each family and each *n*, we drew 10,000 independent sets of *n* embryos without replacement, recorded the minimum *F*_ROH_ in each set, and computed the expected minimum for that cohort size as the average across replicates.

From these quantities we derived the expected reduction in *F*_ROH_ relative to random embryo choice, defined as the difference between family mean *F*_ROH_ and expected minimum *F*_ROH_. Simulations for first-cousin and double-first-cousin couples were analyzed separately and then summarized across the 1,000 families in each group.

Founders were assumed to have no background ROH, so simulated *F*_ROH_ reflects only autozygosity arising from the explicit consanguineous pedigree. We restricted attention to autosomes and did not simulate sex chromosomes. Within these constraints, the simulation captures stochastic segregation of ancestral haplotypes and the resulting variability in *F*_ROH_ among em-bryos of consanguineous unions. All simulations and analyses were performed in Python using custom scripts.

### Estimating risk reductions for monogenic and polygenic outcomes

We modeled risk reductions achieved by combining PGT-M and *F*_ROH_-based embryo selection for outcomes with different genetic architectures. For largely monogenic outcomes (infant death, intellectual disability, congenital anomalies), we assumed that the excess risk above the outbred baseline is entirely attributable to increased recessive burden in consanguineous families. For polygenic outcomes (type 2 diabetes and childlessness) we assumed that log-odds vary approximately linearly with *F*_ROH_ and used effect sizes estimated in within-sibling analyses to minimize confounding from shared environment ^3–5^.

Let *p*_baseline_ denote the lifetime risk in an outbred population (e.g. for type 2 diabetes we used published estimates for the U.S. population ^25^). For a parental relatedness corresponding to *F*_ROH_ = *F*, we let *RR*_*F*_ denote the risk ratio for offspring relative to *F* = 0, estimated from cohort studies of consanguinity and autozygosity ^2–5,7,15,16^. The absolute risk without screening is then

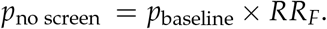

To approximate the impact of PGT-M, we used the diagnostic yield in patient cohorts with the specified condition as a proxy for the fraction of recessive risk attributable to known pathogenic variants. For intellectual disability and congenital anomalies we drew on recent exome sequencing studies in diverse populations ^10,12–14,28^. If *D* denotes the diagnostic yield (e.g. 30 – 50% for severe intellectual disability; 26 – 50% for major congenital anomalies), we modeled PGT-M as removing a fraction *D* of the excess risk above *p*_baseline_ as well as the risk due to biallelic causes in a baseline, *B*, outbred population (assumed to be 4%), yielding

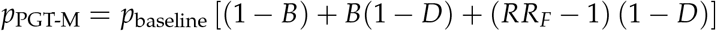

We neglected the slight reduction in *F*_ROH_ that is expected to result from PGT-M, given that PGT-M excludes embryos that are homozygous at the risk locus and thus have on average a slightly higher *F*_ROH_ compared to the other embryos. We note this is likely an overestimate of the achievable PGT-M benefit, because diagnostic yield in affected individuals may include variants that would not meet strict criteria for preimplantation testing. We also assume the PGT-M risk reduction of the baseline risk of biallelic monogenic causes is equal to that of the excess risk due to consanguinity.

Autozygosity-informed embryo selection further reduces risk by lowering *F*_ROH_ from its base-line family value *F* to a post-screening value *F*_post_ when selecting the lowest-*F*_ROH_ embryo among *n* candidates. For largely monogenic outcomes, we assumed that the residual excess risk after PGT-M scales linearly with *F*_ROH_, so that

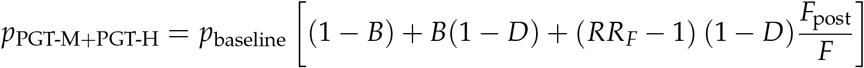

For polygenic outcomes we instead modeled log-odds as proportional to *F*_ROH_ and applied the same *F*_post_/*F* scaling on the logit scale based on within-sibling effect estimates. In both cases, we assumed that recessive contributions to *p*_baseline_ are negligible in outbred populations, consistent with empirical estimates in cohorts with low autozygosity.

Baseline risks for infant death, intellectual disability, congenital anomalies, and type 2 diabetes were drawn from U.S. vital statistics and large epidemiological studies ^22–25^. We used fertility data from the National Center for Family and Marriage Research and the IPUMS Current Population Survey to approximate lifetime childlessness ^26,27^. When multiple sources were available, we averaged lifetime risk estimates across recent, high-quality studies.

## Data Availability

Data supporting the findings of this study are contained within the manuscript and its supplementary materials.

## Supplement: A model for ROH reduction

We follow the classic model of Thomas et al. ^38^ and its recent applications ^39,40^. Denote by *m* the number of meioses from the common ancestor to a hypothetical child of the related couple. Following Figure 1, *m* = 6 when the parents are first cousins or double first cousins, and *m* = 8 for second cousins. The model assumes that recombination chops each chromosome into equal-length blocks. Denote by *L* the total autosomal genome length in Morgan (equal to the number of autosomal recombination events per meiosis) and by *c* = 22 the number of autosomes; the number of blocks is *mL* + *c*. The two copies of each block of the hypothetical child are identical by descent with probability *F*, which is the inbreeding coefficient. We have *F* = 1/16 for first cousins, *F* = 1/8 for double first cousins, and *F* = 1/64 for second cousins. Thus, the number of blocks that would be homozygous (by descent from the recent common ancestor) in the child is approximately a binomial variable, Bin(*mL* + *c, F*). We can approximate *F*_ROH_ as *f*_*H*_, the proportion of homozygous blocks. The variance of *f*_*H*_ is

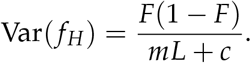

The above expression has two limitations. First, it is an underestimate of the variance of *F*_ROH_, because, in reality, block lengths are variable due to the randomness of recombination. On the other hand, the above expression for Var(*f*_*H*_) includes variability both between-family and within-family, whereas only the within-family variability is of interest for embryo screening. It is thus an overestimate of the within-family variance of *F*_ROH_. Overall, these two deviations partly cancel out, so we use the above expression as an approximation for the within-family variance Var_*w*_(*F*_ROH_).

In previous work (Karavani et al. ^41^), embryo selection for traits was considered, and the mean change in a polygenic trait relative to random selection was derived. Suppose *n* embryos are available. Figure 1 shows that the distribution of *F*_ROH_ within family is approximately normal, similarly to the distribution of a polygenic score. We thus use the result from Karavani et al. Denote by *R* the value of the maximum of *n* standard normal variables. Asymptotically, *E*(*R*) *≈* 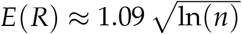. The mean difference in *F*_ROH_ between a randomly selected embryo and the embryo with the lowest *F*_ROH_ is given by

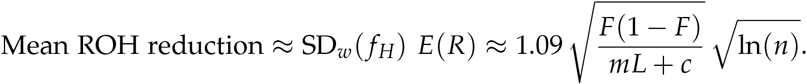

Plugging *L* = 35 M ^42^, we have, for first cousins 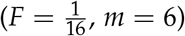:

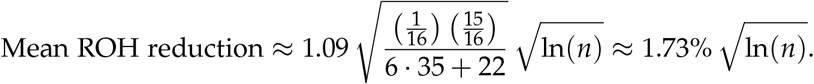

For second cousins 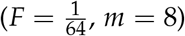:

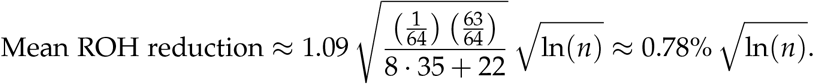

We also consider a second model of within-family *F*_ROH_ variance estimation derived from the simulation framework. Since the distribution has positive skew (Supplementary Figure 1A) and we are interested in approximating the lower half of the distribution as normal, we estimated within-family variance by reflecting the values below the median and estimating variance using this augmented, symmetric distribution.

We then compared the simulated reductions from Figure 3 to the theoretical expectations derived here. Both approaches work well in the case of second cousin parental relatedness (Supplementary Figure 1B). However, in the case of first cousin parental relatedness, the initial model appeared to underestimate the simulated reduction in *F*_ROH_, while the second model provided a relatively good fit (Supplementary Figure 1C). Thus, it appears that modelling risk reduction based on a normal approximation of the *F*_ROH_ distribution was appropriate, though our derivation of within-family variance in *F*_ROH_ is miscalibrated in the case of first cousin parental relatedness.

**Supplementary Figure 1:**
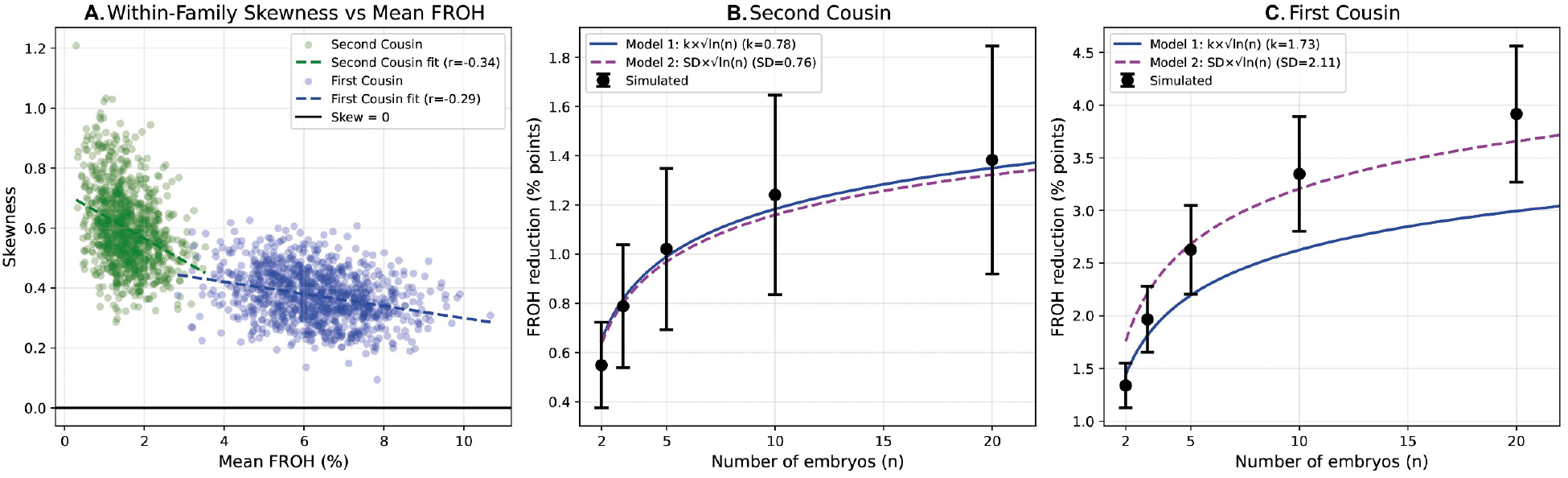
Theoretical expected reduction in embryo ROH from screening in second- and first-cousin unions. (A)Within-family skewness of F_ROH_ distribution versus mean F_ROH_ for 1,000 simulated families from second-cousin (green) and first-cousin (blue) parental matings. Dashed lines show linear fits; horizontal black line indicates zero skewness. (B) Expected absolute reduction in F_ROH_ when selecting the lowest-F_ROH_ embryo from sets of n = 2, 3, 5, 10, 20 embryos for second-cousin parental relatedness. Black points show simulated means ±SD. Model 1 (solid blue): 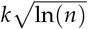 with k = 0.78; Model 2 (dashed magenta): 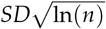 where SD = 0.76 is the symmetric within-family standard deviation. (C) As in (B), for first-cousin parental relatedness, with k = 1.73 and SD = 2.11.

## Footnotes

1 For example, large terminal or chromosome-scale ROH segments that are inconsistent with the expected pedigree structure may warrant clinical review for technical artifacts, uniparental disomy, or other non-Mendelian mechanisms. This quality-control use is distinct from the primary purpose of PGT-H, which is embryo prioritization based on genome-wide realized autozygosity.

